# Left atrial wall shear stress distribution correlates with atrial endocardial electrogram voltage and fibrosis in patients with atrial fibrillation

**DOI:** 10.1101/2024.07.11.24310174

**Authors:** Dionysios Adamopoulos, Georgios Rovas, Nicolas Johner, Hajo Müller, Jean-François Deux, Lindsey A. Crowe, Jean-Paul Vallée, François Mach, Nikolaos Stergiopulos, Dipen Shah

## Abstract

Left atrial (LA) wall fibrosis plays an important role in the perpetuation of atrial fibrillation (AF) since the abnormal electrophysiological properties of the fibrotic areas sustains the arrhythmia by favoring both re-entry circuits as well as abnormal impulse generation. Despite its crucial contribution, the mechanisms by which LA fibrosis develops are not well understood.

The LA wall is constantly exposed to the hydraulic forces exerted by the blood flow arriving from the pulmonary veins. The purpose of the present study was to examine the association between regional wall shear stress and areas with fibrosis in the LA of patients with AF.

15 patients (13 males, mean age 61±11 years) with AF, no significant mitral regurgitation and clinical indication for a primary catheter ablation were prospectively recruited for the study. All participants underwent a baseline three-dimensional electro-anatomical mapping of the LA during the ablation procedure and a pre-interventional cardiovascular magnetic resonance (CMR) imaging with phase contrast for mitral flow estimation and Gadolinium injection for LA fibrosis detection. Fibrotic areas were detected either by low bipolar voltage (BV≤0.5mV) and/or by areas with enhanced late Gadolinium uptake as assessed by the image intensity ratio (IIR≥1.2). For all subjects, a detailed 3D anatomical model of the LA was extracted from the invasive electro-anatomical maps and was used to calculate regional time-averaged wall shear stress (TAWSS) and blood age (BA), an index of blood stagnation, by performing patient-specific computational fluid dynamic simulations.

Globally, areas around the pulmonary veins and the LA roof exhibited the highest values of TAWSS. In all cases, high TAWSS was strongly correlated with low voltage (n=15, r from -0.002 to -0.449, p<0.01) and enhanced late Gadolinium uptake (n=12, r from 0.071 to 0.475, p<0.001). Fibrotic areas as detected by both low BV and high IIR were more prevalent in areas exposed to high TAWSS (21.6% vs 8.1% and 26.2% vs 13.2% respectively, p<0.001). Inversely, in all but one case, areas with low TAWSS presented more intense blood stagnation as assessed by the highest BA (r from -0.268 to - 0.688, p<0.001).

In patients with AF, regional high TAWSS is associated with corresponding CMR biomarkers of left atrial wall fibrosis and electrical scaring. Inversely, areas with low TAWSS are associated with blood stagnation and could favor thrombus formation. This may provide insights of a novel pathophysiological mechanism explaining the characteristic atrial electrical remodeling and thrombus formation seen in patients with AF.

## Introduction

Atrial fibrillation (AF) is the most frequent sustained arrhythmia worldwide, with significant morbidity and mortality rates, especially in the elderly. In the Global Burden of Disease 2010 Study, AF was estimated to affect around 33 million globally, while it was found to be strongly associated with cerebrovascular events, heart failure, hospitalisations, and death(1).

The presence of fibrotic areas in the left atrial wall plays a pivotal role in the perpetuation of AF(2). It is widely accepted nowadays that the pathophysiology of AF comprises two distinct (though equally important) mechanisms: i) the presence of multiple rapidly firing ectopic foci found principally around the pulmonary veins, that act as triggers initiating the arrhythmia(3) and ii) the presence of atrial myocardial cells with impaired electrical conductivity (electrical scars) and/or abnormal repolarization characteristics, that are capable of sustaining the arrhythmia through the formation of pathological electrical wavelets, re-entry circuits and abnormal electrical impulses(4). Although the trigger component of AF has been extensively studied, very little is known about the mechanisms by which electrical scars are created, leading to the perpetuation of the arrhythmia.

Left atrial myocardial cells are constantly exposed to the hydrodynamic forces of the blood flow arriving from the pulmonary veins. In particular, wall shear stress (defined as the frictional force per unit area exerted by the blood flow tangentially on the wall) has been recognized as a major hemodynamic factor affecting both function and geometry in different parts of the cardiovascular system(5). More precisely, the exposure of the arterial wall to pathological (both high and low) shear stress has been consistently associated with progressive vascular damage including atherosclerosis, aneurysm growth and rupture, through both inflammatory-cell and mural-cell mediated pathways(6). Indeed, the exposure of the arterial wall to high wall shear stress has been shown to initiate biochemical cascades (through endothelial cell mechano-transduction) that lead to the local production of proteases and the final apoptosis of the wall’s smooth muscle cells(7).

Based on the above-mentioned observation, we hypothesized that the exposure of the left atrial wall to high shear stress may have similar effects leading to cell apoptosis and thus contributing to the formation of fibrotic areas in the left atrial wall. The aim of the present study was to examine the association between regional wall shear stress and fibrotic areas exhibiting impaired endocardial voltages in the left atrial wall of patients with AF.

## Materials and Methods

### i) Study population

15 patients (13 males, mean age 61±11 years) were prospectively recruited for the study. All presented symptomatic episodes of AF with a clinical indication for catheter ablation. 10 subjects presented with paroxysmal and 5 with persistent AF, while 3 presented additionally atrial flutter. All patients underwent a baseline echocardiography and only patients without hemodynamically significant (more than moderate) mitral regurgitation were eligible for the study. Informed written consent was obtained from each patient and data were anonymized prior to analysis. The Commission Cantonale d’Éthique de la Recherche sur l’être humain of the Canton of Geneva gave ethical approval for this work, approval number: 2023-02314.

### ii) Cardiovascular magnetic resonance imaging (CMR)

#### a) CMR acquisition protocol

All patients underwent a CMR at baseline before the catheter ablation (5±3 days before the ablation, range same day to 8 days before). All scans were performed using a 3 T clinical MRI system (IMRIS (IMRIS, Deerfield Imaging, Minnetonka, MN) ceiling-mounted iMRI System combined with Artis electrophysiology suite (Siemens Erlangen, DE, MAGNETOM Skyra/Artis)) using two 18-channel array coils (anterior and posterior). The CMR protocol included balanced steady-state free precession (SSFP) cine imaging in long-axis orientation (two-chamber, four-chamber and short axis stack views). An ECG gated free-breathing 3D contrast-enhanced MR angiogram of the left atrium and the pulmonary veins was obtained immediately after injection of contrast agent (DotaremVR, Guerbet, Roissy, France). For the left atrial fibrosis, a 3D late enhancement sequence (LGE-CMR) was acquired 15 minutes later (Siemens prototype ‘Whole-Heart’) with Dixon fat suppression and 100% efficiency iNav respiratory motion navigator during a static period of the cardiac cycle in terms of atrial contraction. The typical acquisition parameters were: repetition time10 ms; TE1 and TE2 1.3, 2.8 ms; flip angle, 25°; in-plane resolution 1.3×1.3mm with slice thickness 1.3mm; acquisition time 7 min 52 +/-1 min 46. For the phase contrast flow quantification acquisition for the mitral valve parameters were TR/TE 28/2.4 ms; in-plane resolution 1.9×1.9mm with slice thickness 6mm; velocity encoding 250 cm/s; grappa acceleration factor 2; acquisition time 116 heart beats.

#### b) CMR image analysis

The CMR were obtained during ongoing atrial fibrillation in 2 and during sinus rhythm in 13 subjects. Segmentation of the left atrium was performed in a free, open-source software for image analysis (3D slicer)(8). Initially, the blood pool of the left atrium (including the pulmonary veins) was manually segmented on the CMR axial slices. A two-voxel surface dilation was used for delimiting the epicardial border, and thus a 3D model of the left atrial wall was created. The mitral annulus was used to separate the left atrium from the left ventricular cavity. On the 3D LGE-CMR images, the signal intensity was normalized to the mean blood pool intensity as described in the image intensity ratio (IIR) method(9, 10). CMR image analysis was not performed for 3 subjects due to technical issues with image acquisition.

### iii) Catheter ablation

Catheter ablation procedures were performed following institutional standard of care. Transfemoral, transseptal access was obtained and left atrial pressures were recorded. A 10-electrode catheter (Inquiry, Abbott, Abbott Park, Illinois, USA) was placed in the distal coronary sinus or great cardiac vein and a second similar 10 electrode catheter was placed with its proximal electrodes at the high right atrium and its tip near the lateral cavotricuspid isthmus.

In patients who presented in sinus rhythm, a standardized AF induction protocol was performed using right atrial decremental burst pacing with 8-beat sequences, starting with a 300 ms pacing cycle length down to 200 ms (or effective atrial refractory period) by steps of 10 ms.

A 3D electro-anatomical map (CARTO mapping system, Biosense Webster, Irvine, California, USA) of the left atrium was obtained in AF using a 20-electrode circular mapping catheter (LASSO Catheter, Biosense Webster). For 8 patients the map was obtained in sinus rhythm because sustained AF was not inducible or not induced. Complete left atrial anatomy (including proximal pulmonary veins) was obtained by fast anatomical mapping (FAM, Biosense Webster) and a bipolar voltage (BV) map was obtained by measuring the maximum bipolar peak to peak voltage amplitude during the annotation window.

In all subjects, circumferential pulmonary vein isolation was performed (Thermocool SmartTouch SF Catheter, Biosense Webster), using a power range of 25-35W and application duration of 20-30 seconds titrated following standard practice. Pulmonary vein isolation (entrance block) was confirmed using the circular mapping catheter. Extra-pulmonary vein substrate modification was performed at operator’s discretion, targeting fractionated electrograms in the coronary sinus, left atrium, and/or right atrium. Additionally, linear ablation lesions (e.g., left atrial roof) were performed at operator’s discretion based on substrate distribution. Bipolar electrograms at baseline were filtered (band-pass 30-500 Hz) and digitally recorded at a sampling frequency of 1,000 Hz (LabSystem Pro, Boston Scientific, Marlborough, Massachusetts, USA) along with surface ECG.

### iv) Computational fluid dynamics (CFD)

The coordinates and measured variables of all data points recorded during the catheter ablation procedure were extracted and were used to generate a patient-specific 3D model by utilizing the Poisson surface reconstruction (Figure 1). Flow extensions were added on the pulmonary veins and the mitral valve with a length equal to ten times the equivalent diameter of each orifice to ensure that the flow is fully developed. The 3D model with the flow extensions was meshed with polyhedral elements and a five-element-wide prismatic boundary layer on all wall surfaces. The meshing parameters were kept constants for all cases, resulting in volumetric mesh sizes in the range of 0.9–1.7 million elements, depending on the size of the left atrium.

**Figure 1.**
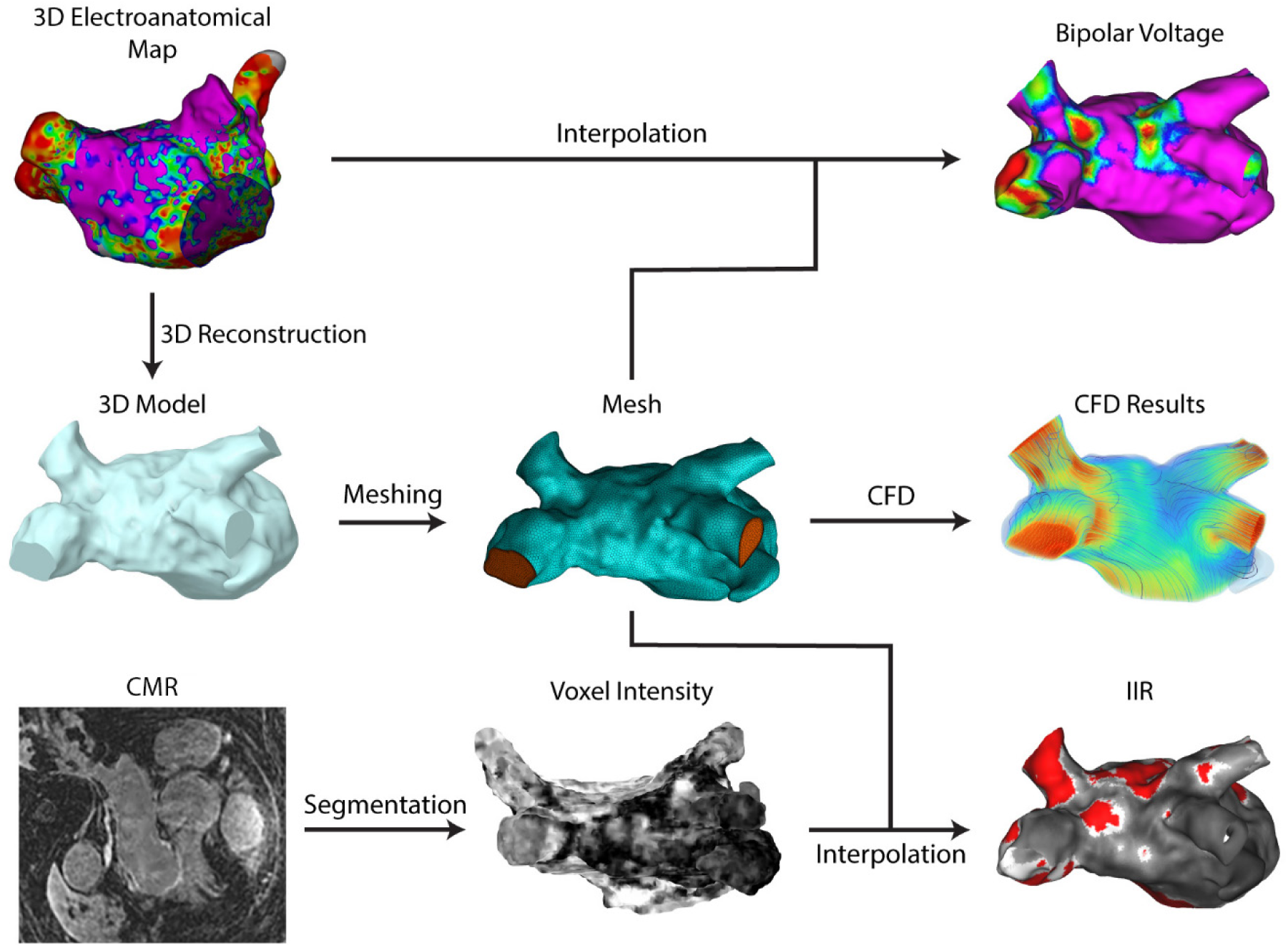
Outline of the methodological steps, integrating data from the electro-anatomical maps (left atrium geometry and bipolar voltage), computational fluid dynamic (CFD) analysis and CMR with Gadolinium contrast (IIR: image intensity ratio).

We performed a transient CFD simulation by running the model for six consecutive heart cycles to allow for the stabilization of the transient phenomena. The time step was fixed at 0.25 ms, which was sufficient for the solution to converge in all cases and time steps. The flow was considered laminar, an assumption that was verified by checking the maximum Reynolds number. The blood was modeled as a Carreau-Yasuda fluid(11). We imposed a patient-specific periodic velocity profile at the inlets, which was derived from the phase contrast CMR (Figure 2.a, b). This constitutes a good assumption when individual data for each pulmonary vein are not available, as was proven by a similar computational analysis(12). The measured profile also controlled the period of the simulation. At the outlet, we imposed the mean atrial pressure, as measured during the ablation procedure. The walls were assumed rigid, due to the limited wall motion that occurs during AF(11). A diffusion equation was added to the flow equations and a source term was added throughout the atrium, to calculate the distribution of the blood age (BA) throughout the atrial domain, as described previously(13). The distribution of BA was measured at a distance of 1 mm from the atrial wall, to avoid wall boundary effects. Second-order discretization schemes were selected for all the model variables. The models were solved in the High-Performance Clusters of EPFL (SCITAS, Lausanne, Switzerland), while the average computational time for each case was 30 hours. The meshing and the simulation were performed in Fluent (Ansys, Canonsburg, PA).

**Figure 2.**
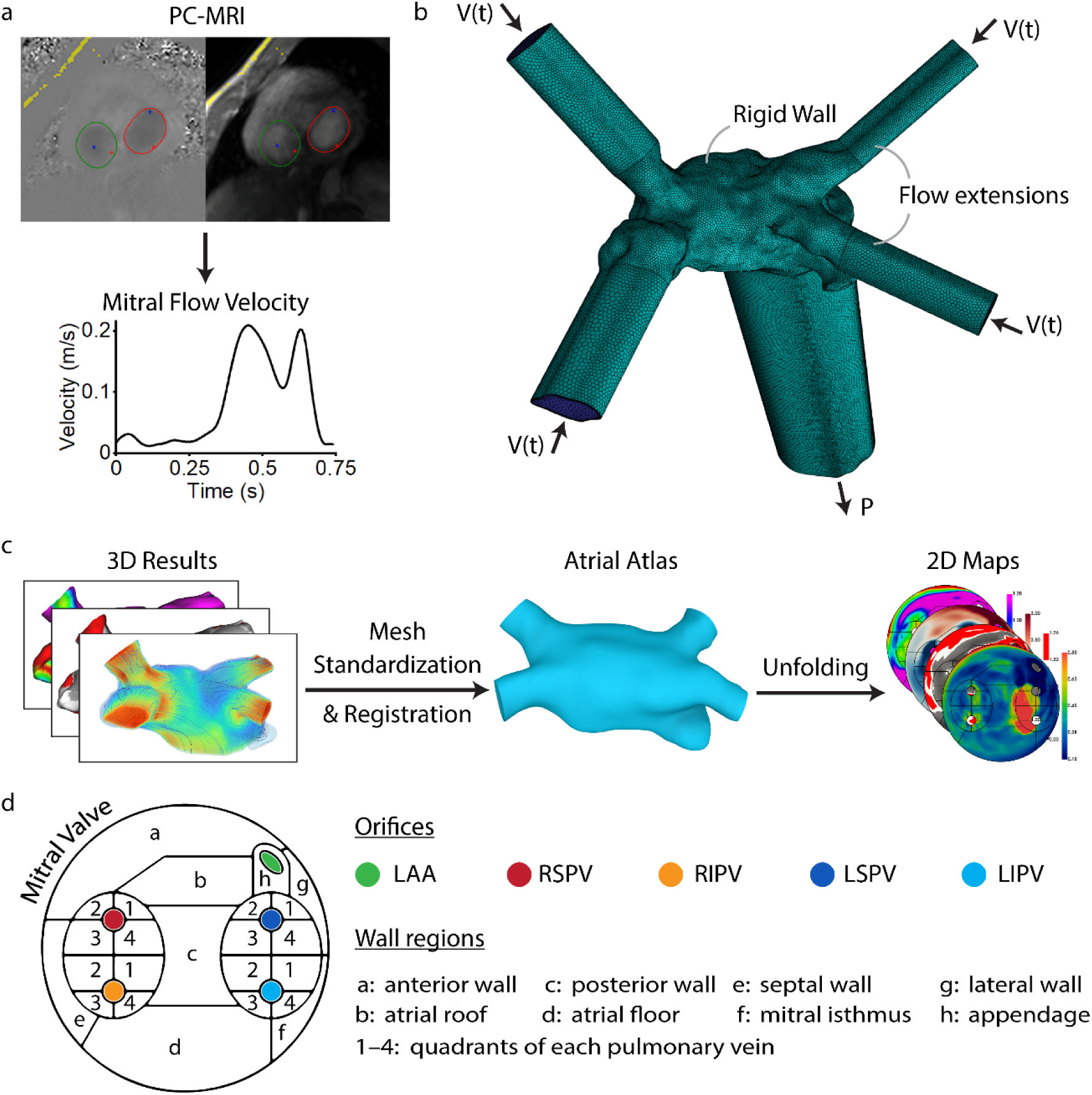
CFD and post-processing methodological overview. a) Derivation of patient-specific mitral flow velocity from phase-contrast CMR (PC-CMR). b) Example of the boundary conditions and flow extensions on a 3D mesh. c) The methodological steps of the unfolding procedure to generate the 2D maps via a standardized atrial atlas. d) The regions of the 2D atrial maps. V(t): time-dependent flow velocity, P: pressure, LAA: left atrial appendage, RSPV: right superior pulmonary vein, RIPV: right inferior pulmonary vein, LSPV: left superior pulmonary vein, LIPV: left inferior pulmonary vein.

### v) Data post-processing

We exported the flow variables of the last heart cycle of each CFD simulation. We used the exported time-dependent distribution of wall shear stress on the atrial wall surface to calculate common indices of shear stress, following the standard methodology(14, 15). Specifically, we calculated the time-averaged wall shear stress (TAWSS), the oscillatory shear index (OSI), the endothelial cell activation potential (ECAP), the relative residence time (RRT), the highly oscillatory, low-magnitude shear (HOLMES) index and the time-averaged wall shear stress gradient (WSSG) on the flow direction as follows:

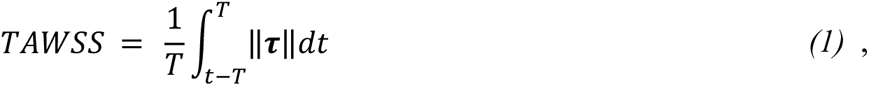

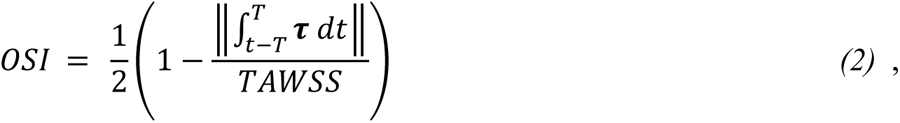

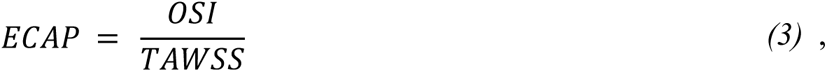

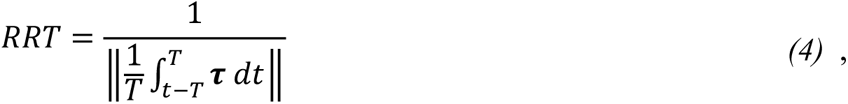

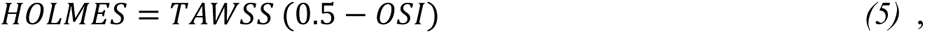

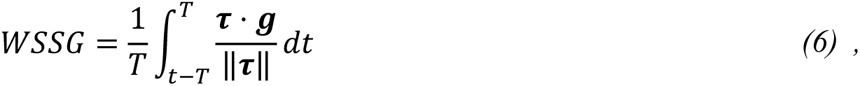

where T is the heart period, t is the time, **τ** is the vector of the wall shear stress and ***g*** is the vector of the projection of the WSS gradient of the flow direction(16). These wall shear indices have been developed initially for computational studies on atherosclerosis and aneurysms, but they have shown great precision in localizing regions prone to structural and functional changes. The combination of high wall shear stress and positive WSSG has been shown to induce internal elastic lamina damage, aneurysm formation and mural-cell-mediated remodeling, while low WSS combined with high OSI can cause inflammatory response on the aneurysmal wall(6, 16, 17). The HOLMES index was proposed to quantify in a single index the combined effect of low but highly oscillatory wall shear(18). Similarly, ECAP was suggested as an alternative index to localize regions with high OSI and low WSS by quantifying the susceptibility to thrombus formation(19). The RRT is strongly correlated with OSI and serves as an index of the near-wall blood stagnation(20), but, despite its name, it remains a wall shear index that differs from BA, which is directly measured via the previously described methodology.

Consequently, we transferred the measured values of BV and IIR on the atrial wall surface used for the CFD simulation, to allow for quantitative comparison among the measured and calculated variables (Figure 1). This was achieved by first aligning the CMR segmentation with the CFD surface mesh, using an Iterative Closest Point algorithm, and then by interpolating the IIR and BV values on the CFD mesh utilizing 3D Radial Basis Function interpolation. The choice of the CFD mesh as the basis of this transform was because most variables had already been calculated on it. This procedure resulted in a set of points with their corresponding BV, IIR and wall shear indices for each subject and these values were used for the subsequent statistical analysis.

To create the two-dimensional (2D) maps, the CFD mesh was standardized by clipping the pulmonary veins to a fixed length of 10 mm, and then it was registered to an anatomical atlas of the left atrium (Figure 2.c). The registered mesh was unfolded by constraining the mitral valve to the perimeter of a predefined 2D pattern to create the 2D maps of every variable and automatically separate the atrial wall in 24 regions (Figure 2.d). The unfolding process has been previously described in detail(21), while the 2D maps serve as an equivalent of the standardized bullseye diagrams that are used for the left ventricle. We calculated the mean value of each variable per region of 2D atrial map *x̄*_*i*_ as the arithmetic mean, and the normalized mean value per region as 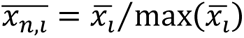, where x is the variable, x_n_ the normalized variable and i the region number. The post-processing pipeline was automated, by requiring minimal user input and was performed using in-house algorithms programmed in Python (Python Software Foundation, Wilmington, DE) and Matlab (MathWorks, Natick, MA). These algorithms are available on https://github.com/g-rov/lausm.

### vi) Statistical Analysis

Categorical variables are expressed as percentages. Continuous variables are expressed as means ± standard deviation, unless differently mentioned. Normality was assessed by visual inspection of the frequency distributions for each continuous variable. Correlations between shear stress indices, BV and IIR were assessed individually for each case using the Pearson correlation coefficient. TAWSS values for each case were further grouped into quartiles (Q1-Q4). Comparisons among groups for categorical variables were performed by using Chi-square test. For continuous variables comparisons were performed after analysis of variance (ANOVA). Levene’s test was used to assess the homogeneity of variance among the compared groups, and in case of violation, Welch’s ANOVA test was used. The Fisher z-transformation was applied to the correlation coefficients to allow comparison with ANOVA. Statistical significance was assumed at a 2-sided p-value level of 0.05. Statistical analysis was performed in IBM SPSS statistics (IBM Corp. Released 2020. IBM SPSS Statistics for Windows, Version 27.0. Armonk, NY: IBM Corp).

## Results

### i) Regional distribution of TAWSS

The absolute values of TAWSS varied among subjects depending on the size and anatomy of each atrium and the cardiac output. The distribution of TAWSS is illustrated on a 3D model for an exemplary case in Figure 3, and for all cases in the 2D unfolded maps in Supplementary Figure 3. The pulmonary veins had significantly higher TAWSS than the rest of the atrium (p = 0.003). In all cases at least one high TAWSS region was observed near the pulmonary vein ostia. Interestingly, the left pulmonary veins had 22% higher TAWSS compared to the right ones (p < 0.001).

**Figure 3.**
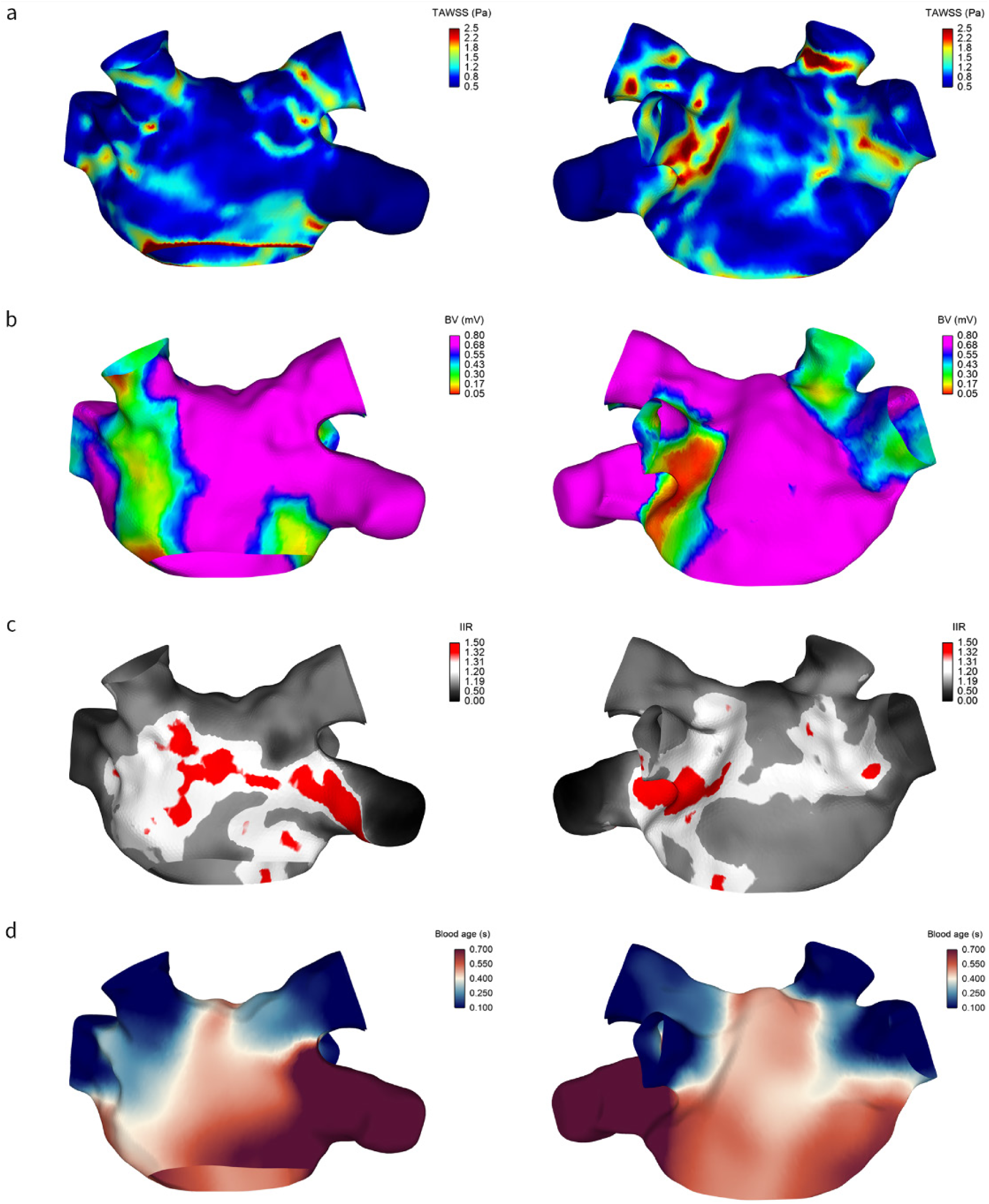
The distribution of wall shear and fibrosis on the 3D atrial model of subject 10 in antero-posterior (left) and postero-anterior (right) views. The colors represent the: a) time-average wall shear stress (TAWSS), b) bipolar voltage (BV), c) image intensity ratio (IIR) separated in regions with fibrosis (red), interstitial fibrosis (white) and no fibrosis (grey color), and d) the blood age (BA).

### ii) Regional distribution of Fibrosis markers

A representative case of the 3D distribution of the electrical scar and CMR fibrosis on the atrial wall is presented in Figure 3. The 2D atrial maps of BV and IIR for all subjects are demonstrated in Supplementary Figure 3. In most of the cases, areas with lower BV were more frequently observed at the roof of the atrium and the area around the pulmonary veins (Supplementary Figure 3). In most of the cases (n=9) a negative association was observed between BV and IIR (Supplementary Figures 1 & 2). Interestingly the inverse relationship was noticed in 3 cases with very low (subject 2 and subject 11) and very high (subject 6) levels of CMR fibrosis (Supplementary Figure 1 & 2).

### iii) Hemodynamics / Fibrosis interplay

There was a notable overlap in regions with low BV, elevated TAWSS and high IIR, as can been seen for four representative cases (Figure 4). TAWSS was found to have a significant negative correlation with BV in all cases (n=15, Pearson r range from -0.021 to -0.449, case 11: p = 0.003, all other cases: p < 0.001) and a significant positive correlation with IIR (n=12, Pearson r range from 0.071 to 0.475, all cases: p < 0.001). The acquisition of the 3D electro-anatomical map in sinus rhythm or in AF did not significantly affect the correlation coefficient of BV with TAWSS (p = 0.20) or with BA (p = 0.14). Regarding their correlation with fibrosis/electrical scar, the other shear stress indices that we evaluated had: i) non-significant correlations, ii) good correlation with only one of the two measures of fibrosis, iii) lower correlations compared to TAWSS, or iv) inconsistent (both positive and negative) correlations, depending on the subject (Supplementary Figure 1 & 2).

**Figure 4.**
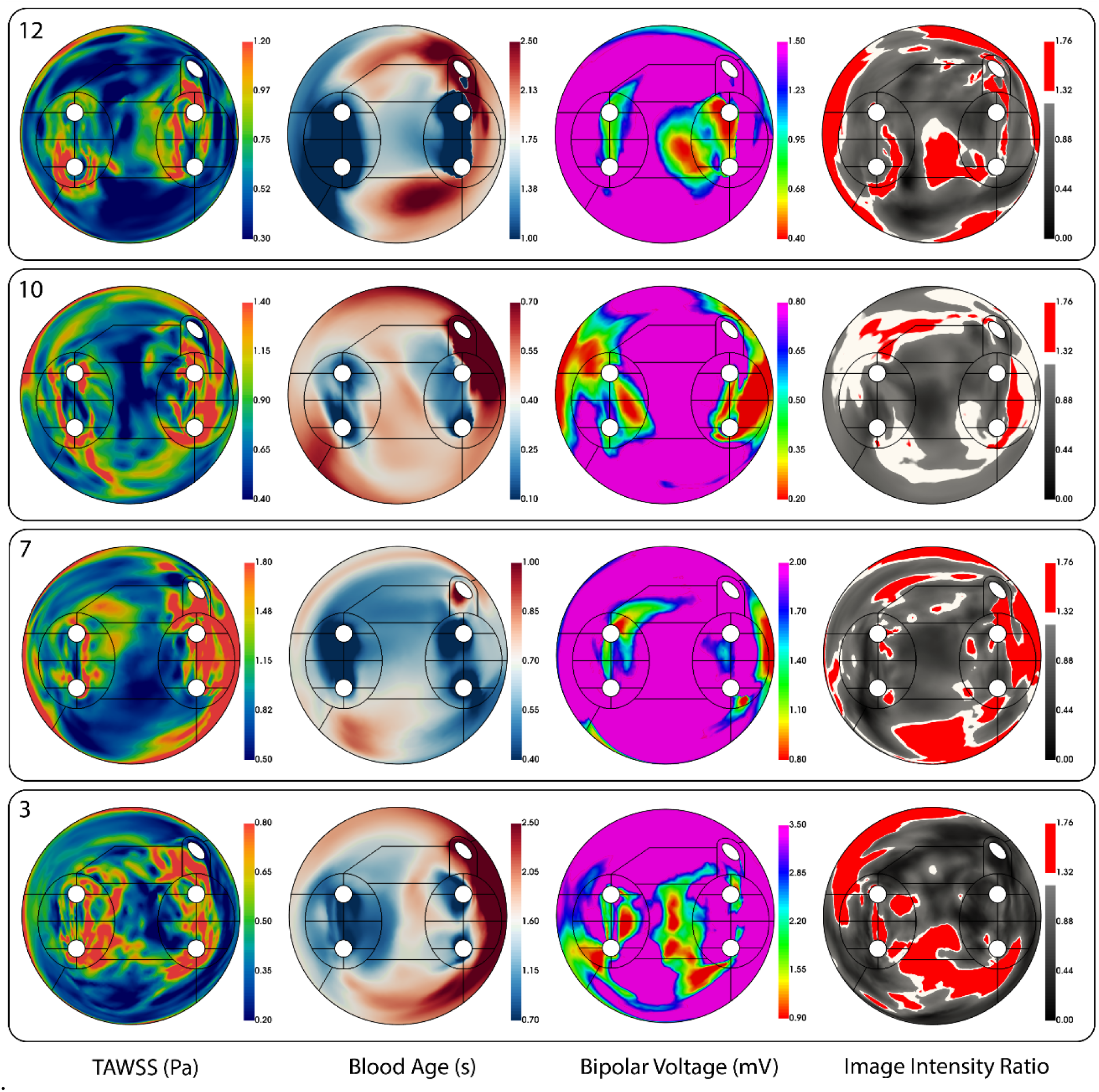
The combined 2D maps of four representative subjects of the primary variables: Time-averaged wall shear stress (TAWSS), Blood age, Bipolar Voltage and Image intensity ratio. Each panel corresponds to one subject denoted by the corresponding subject number.

To further quantify the relationship between wall shear stress and fibrosis, we separated the TAWSS values into four quartiles and calculated the BV and IIR of each quartile for each subject (Figure 5). We observed a consistent trend for both BV and IIR with increasing TAWSS in all cases, although to a different extent. The means of BV and IIR of each quartile were found to be significantly different in all cases (p < 0.001). Finally, regions with higher TAWSS exhibited higher degree of fibrosis/electric scar as detected by both low BV and high IIR according to the standard diagnostic criteria for each method(10, 22) (Figure 6, p<0.001).

**Figure 5.**
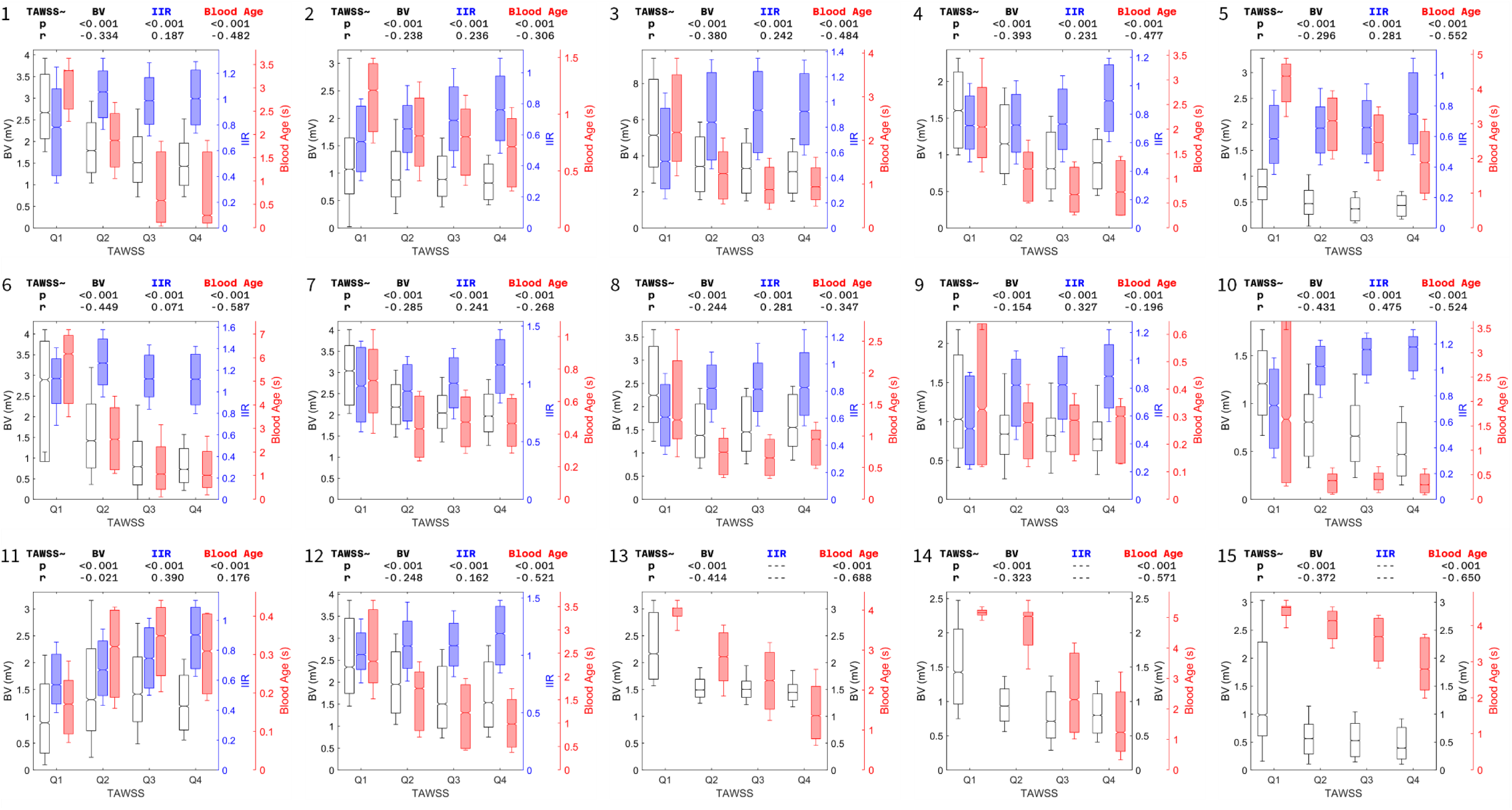
The interplay between time-average wall shear stress (TAWSS) and fibrosis, as measured via the bipolar voltage (BV), the image intensity ratio (IIR) and the blood age of each subject. The TAWSS is divided into quartiles (Q1–Q4). On top of each chart is the corresponding p-value of the ANOVA between the quartiles of TAWSS and BV, IIR or Blood age, and the Pearson’s correlation coefficient (r) between all data points of the same variable pairs. The boxes correspond to the interquartile range, the notches to the 95% confidence interval of the median and the whiskers to the standard deviation. LGE-CMR acquisitions were not available for subjects 13–15.

**Figure 6.**
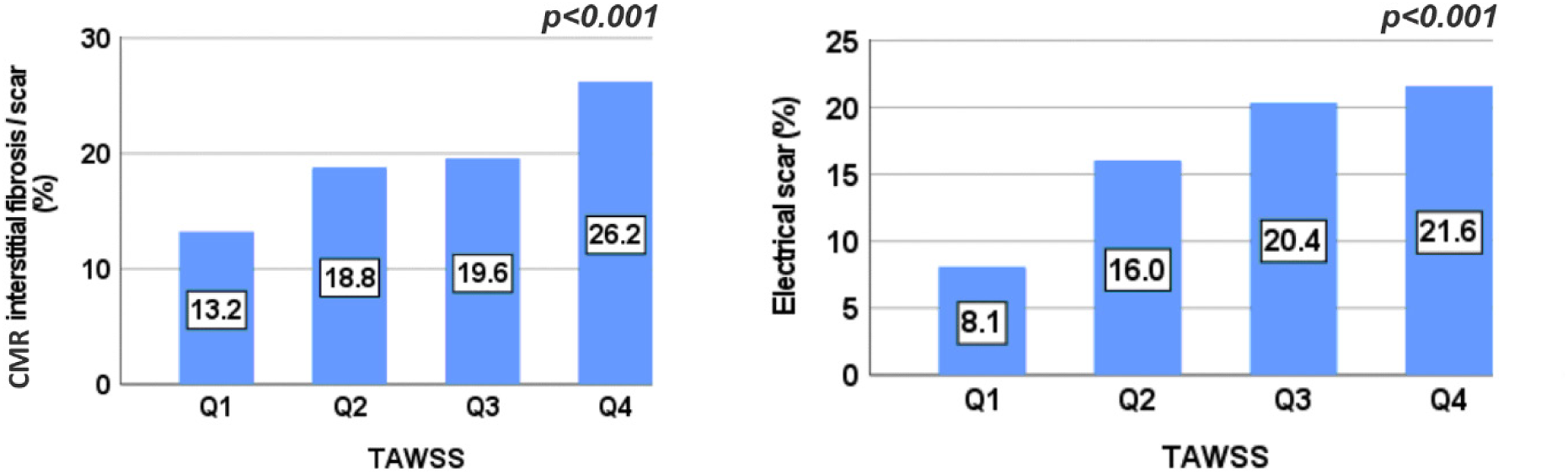
CMR derived fibrosis (IIR ≥ 1.2) and electrical scar (BV < 0.5mV) according to the TAWSS quartiles for the total study population.

### iv) Regional distribution of blood age

Differences in the regional distribution of BA were noted in all study participants (figure 7, p<0.001). The longest BA duration was observed consistently in the left atrium appendage with shortest values in the pulmonary veins and at their ostial junction with the left atrium. A point-to-point comparison between TAWSS and BA showed strong, inverse correlations in all but one case (Pearson r range from -0.268 to -0.688, p<0.001 for all, exception case 11, Figure 5, Supplementary Figure 3). BA also showed strong positive correlation with BV in all cases (Pearson r range from 0.24 to 0.78, p<0.001 for all, Supplementary Figure 1).

**Figure 7.**
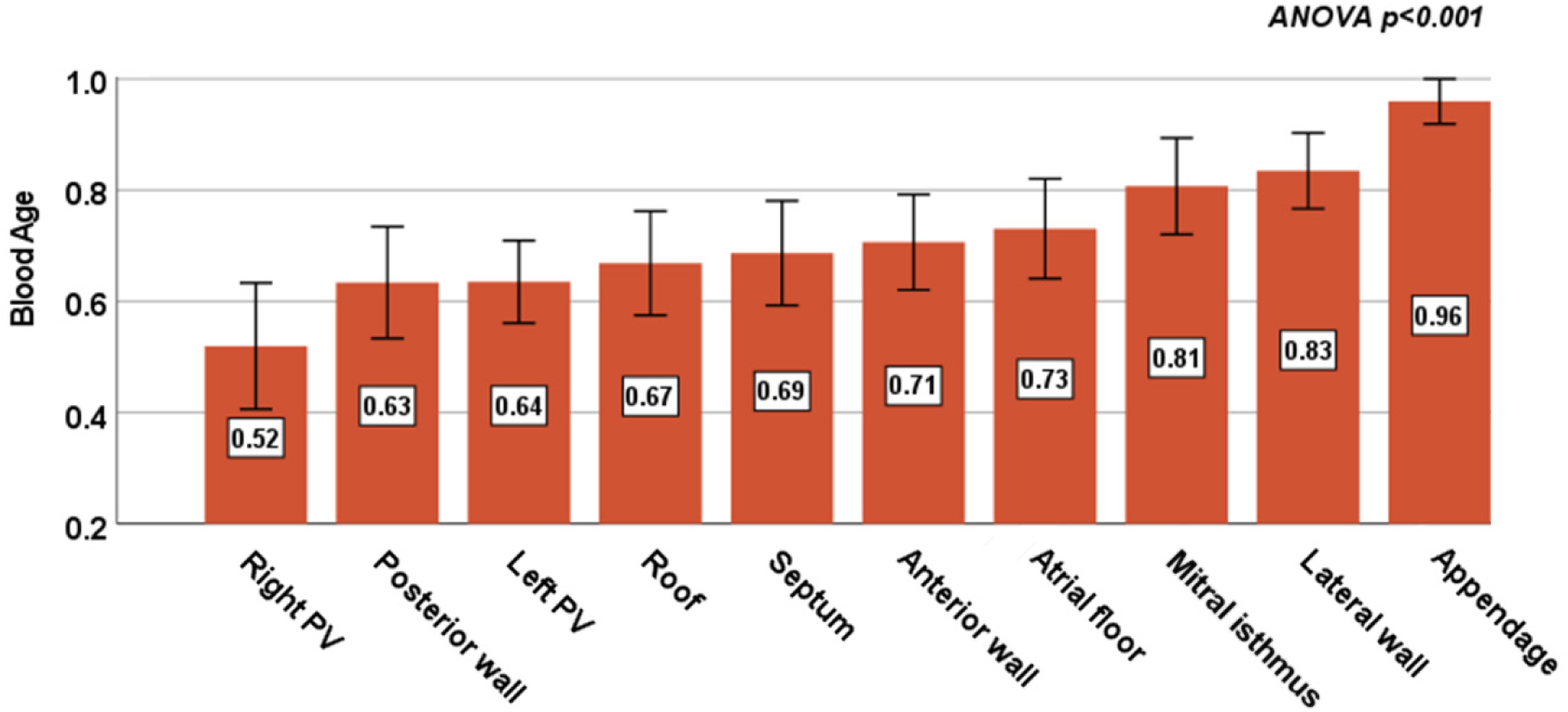
Normalized blood age (BA) according to the left atrium regions for the total study population. PV: Pulmonary veins. The error bars indicate the 95% CI.

## Discussion

To the best of our knowledge, this is the first attempt to associate hemodynamic indices, specifically wall shear stress, with atrial wall fibrosis as assessed by both electrical mapping and CMR imaging. The main findings of the present study can be summarized as follows: a) in patients with AF the regional distribution of fibrosis/electrical scar as assessed by both BV and IIR follows specific patterns with an important presence in the areas around the pulmonary veins and the roof of the left atrium, b) the same regional distribution is also observed for wall shear stress, which shows significant correlations with the fibrosis/electrical scar indices. Consequently, in patients with AF, areas with high wall shear stress exhibit more pronounced fibrosis/electrical scarring, pointing to a novel pathophysiological link explaining the well-established association between AF and pathologies with an impaired left atrium hemodynamic environment (e.g. systolic or diastolic left ventricular dysfunction, mitral regurgitation)(23, 24). c) Inversely, areas with low wall shear stress (e.g. the LA appendage) exhibited the highest blood age denoting a significant link between wall shear stress and conditions favoring thrombus formation.

Many in vitro studies have focused on the cellular response of the atrial myocytes to wall shear stress. Boycott et al. reported that the application of shear stress on atrial myocytes generated an outward current mediated by K^+^ channels and resulted in shortening of the action potential duration. The shear stress threshold for this shear-induced activation was reported at 0.28 Pa, while dilated and fibrotic atria were shown to be less responsive to the shear-induced current increase, indicating that the shear stress could primarily affect the early stages of AF(25). Son et al. showed that shear stress above 0.2 Pa can induce a shear stress-sensitive current on the membrane of atrial myocytes and trigger the release of Ca^2+^ in the subsarcolemmal domains(26). In addition, Yamamoto et al. demonstrated that the endothelial cells react to wall shear stress via a mechano-transduction pathway. Specifically, it was shown that wall shear stress as low as 0.1 Pa is enough to increase ATP production in the mitochondria, followed by increased extracellular ATP release which increases the influx of extracellular Ca^2+^(27). Ca^2+^ release or influx favors abnormal impulse generation, likely as triggered activity. Thus, these findings could well account for electrical remodeling including shortened action potential durations as well as AF initiating triggers. In other observations, Morel et al. showed that high wall shear stress (8 Pa) affected the shape and architecture of endothelial cells, up-regulated the expression of proteins related to cytoskeleton and down-regulated extracellular matrix proteins(28). In our CFD simulations the maximum wall shear stress reached this threshold in all cases, while it ranged up to one order of magnitude higher in certain subjects. Finally, Meng et al showed that high wall shear stress in combination with high WSSG can lead to endothelial cell damage and turnover, extracellular matrix degradation and mural cell apoptosis through cellular mediated mechanisms(6). These pathways provide a molecular background explaining the association between fibrotic remodeling and high wall shear stress observed in our study.

The dependence of the degree of electrical scar and fibrosis on TAWSS appeared to be non-linear, by rapidly increasing with increasing TAWSS and then plateauing (Figure 5). Therefore, the Pearson’s linear correlation coefficient could not accurately summarize this behavior but served more as an indication of this relationship and prompted us to perform subsequent analyses in TAWSS quartiles.

TAWSS achieved good localization of affected regions, as can be confirmed by the visual comparison of the 2D maps especially for the BV (Figure 4, Supplementary Figure 3). The same is true for BA and BV, which was expected given the direct connection of both TAWSS and BA with the velocity field which is derived through the fluid dynamics equations. The fibrotic patterns, as evidenced by the BV maps are replicated with good accuracy in the patterns of TAWSS, even in cases where the statistical analysis does not clearly show this relationship (e.g. case 11). In cases where there is high asymmetry of fibrosis between the left and right side of the left atrium, e.g. cases 8 and 14 which both show increased fibrosis between the left superior and inferior pulmonary veins, the TAWSS results confirm the observed asymmetry. This is of major practical importance since CMR-based estimations of TAWSS become readily available in the clinical routine, providing non-invasive information about potential areas/targets for ablation.

In the present study, CMR derived fibrosis and areas with low endocardial voltage showed different levels of correlation, with the vast majority being negative (n=9 [75%], Supplementary Figures 1 & 2). Atrial fibrosis has been proposed as the main mechanism underlying low bipolar voltage since it interrupts fiber bundle continuity which leads to local conduction abnormalities(29). However, point-by-point data assessing the agreement between LGE-CMR atrial wall intensity and BV are scarce. Harisson et al., found only weak associations between atrial wall signal intensity and endocardial voltage in patients undergoing re-do procedures(30). In a recent study, LGE-CMR derived atrial fibrosis as assessed by the IIR correlated with endocardial BV, with the relationship becoming weaker with atrial dilation(31). These discrepancies may be at least partially explained by technical artefacts and inter-individual variability. Moreover, it has been reported that BV depends on the electrophysiological conditions including a strong dependance on atrial frequency, rhythm and direction of activation waves (32). Another possible explanation for the weaker correlation between atrial wall signal intensity and endocardial voltage may be that early-stage interstitial fibrosis is not detected by the LGE-CMR sequences(33).

Despite the good agreement between TAWSS and fibrotic patterns near the pulmonary veins and the posterior wall, not all fibrotic patches conform to regions with high TAWSS. A common exception are large fibrotic patches on the anterior wall and the atrial floor near the mitral valve, which are not matched by equally large high TAWSS areas. This could be attributed to the effect of outgoing flow through the mitral valve on the local hemodynamics and the absence of a valve leaflet motion model in our simulations, while similar exceptions have been also recorded in previous studies(34).

Consistent patterns of regional distribution of the BA were also noted in all subjects with the appendage presenting the highest blood stagnation index followed by the lateral wall, mitral isthmus, and atrial floor. On the contrary, areas around the ostia of the pulmonary veins, the posterior wall and the atrial roof presented lower blood age (figure 7). As expected, the association with TAWSS is inversed since high velocities contribute to the rapid “cleansing” of the blood from the different areas of the left atrium, whereas low velocities result in blood stagnation. This observation highlights even more the utility of the left atrium TAWSS evaluation since it predicts not only the location of the AF substrate (highest end values) but also areas with high thrombogenic potential (lowest end values). This may contribute to a patient-specific thrombogenic risk assessment that extends beyond classic risk scores in the era of personalized medicine.

Compared to other cardiovascular conditions, the use of CFD, structural or combined fluid-structure interaction simulations has been limited in the case of AF, in terms of both the overall number of studies and the number of patients included in each of them(35). While computational approaches are gaining momentum as an additional diagnostic tool, their results in the left atrium and ventricle have been validated against 4D Flow CMR(12) and certain cardiac CFD methods have even received clinical approval(35). Hunter et al. showed that regions with low voltage and electrical scar have higher wall stress, via structural left atrium models imposed solely to the transmural pressure without accounting for the hemodynamics(36). CFD simulations have been previously used to calculate indices of wall shear stress on the left atrial wall in the presence or absence of AF(37) and to assess catheter ablation outcomes and risk of thrombosis(38, 39), but in none of those cases have the results been compared to measurements of markers of fibrosis. Our results agree well with the reported ranges of hemodynamic indices in the left atrium of patients with AF in those studies.

Wall shear stress indices have been compared in fibrotic and non-fibrotic left atrium regions of patients with AF(34). Although the reported ranges of the shear indices are similar to those presented here, our findings disagree with the distribution of those indices in healthy and fibrotic regions. This discrepancy could be attributed to certain methodological differences as in the previous study the authors used: i) CMR images to derive the CFD model, which has lower spatial resolution and results in smoother model surfaces, ii) generic boundary conditions and generic wall motion, iii) only CMR to identify fibrotic areas, which has lower resolution and specificity compared to BV, and iv) lower number of patients. Furthermore, due to the previous study’s limitation of providing solely summarized data, without the regional distribution of shear indices on the left atrial walls, it hindered the possibility of conducting more comprehensive comparisons aimed at identifying additional reasons for the observed differences.

## Conclusions

We conclude that left atrial wall regions with CMR markers of fibrosis and low endocardial voltage significantly overlap with regions of increased wall shear stress and they develop preferably near the pulmonary veins and the atrial roof. Inversely, areas with low wall shear stress were prone to blood stagnation. These results are supported by patient-specific simulation of the hemodynamic conditions in the left atrium, and they suggest a potential pathophysiological link between AF and disturbed hemodynamic environment. Further developments in CMR-based wall shear measurement and automated CFD workflows of the left atrium could enable the noninvasive localization of regions prone to AF substrate development and regional stasis.

## Limitations

In our study, the wall shear stress is estimated based on CFD methods and has not been measured directly. The CFD methodology is based on assumptions and the application of boundary conditions that may significantly affect the results, although to a small extent. Due to the selected CFD methodology, we decided to exclude patients with significant mitral regurgitation, since the CFD model would be unable to accurately capture regurgitation jets, which could alter the atrial hemodynamics. Secondly, the atrial wall displacement and its interaction with the blood have not been considered in our CFD simulations. The effects of fixed or moving walls on the left atrium hemodynamics have been studied in the past(40). The authors compared the influence of fixed-wall and moving-wall models, albeit with slightly different inlet boundary conditions, on atrial flow patterns, blood residence time and kinetic energy, but not on indices of wall shear. They concluded that incorporation of wall motion affected mostly the flow fields of atria with normal contraction and to a lesser extent fibrillating atria, due to the limited and difficult to model wall motion in the latter case. Considering these findings and due to the absence of patient-specific time-dependent imaging data, we decided to model the wall as stationary in our simulations. Moreover, although results are statistically significant and the conclusions clinically relevant, the small sample size should be acknowledged as a limiting factor. Eight patients in our study did undergo BV mapping in sinus rhythm and the remaining during AF, however, a good correlation has been reported between BV measured in sinus rhythm versus during AF(41). Pertinently, a sensitivity analysis in our cohort showed that the correlation of BV and TAWSS/BA was maintained and similar irrespective of whether the electro-anatomic map was acquired during AF or sinus rhythm. In addition, the results may not be applicable to the general population since specific inclusion criteria and CMR demands were applied. Finally, although most of the processes were semi-automated, operator-independent, segmentation still needed to be manual.

## Data Availability

All data used in this study are available in the Manuscript and the Supplementary Material. The software is available at https://github.com/g-rov/lausm.

## Acknowledgements

We extend our sincere gratitude to Mrs Michaela Schmidt and Mr Karl Kunze (Siemens Healthineers) for the development of the CMR acquisition protocol. We also acknowledge Mr Noé Schmutz and Mrs Charlotte Vivet (Biosense Webster) for the preparation and extraction of the electro-anatomical data.

## Project contribution

The authors confirm contribution to this work as follows: study conception and design: D. Adamopoulos, G. Rovas, D. Shah, F. Mach and N. Stergiopulos; data collection: D. Adamopoulos, D. Shah, N. Johner, JF Deux, LA Crowe, JP Vallée, M Schmidt; analysis and interpretation of the results: D. Adamopoulos, G. Rovas, N. Johner, D. Shah, H. Müller, N. Stergiopulos; draft manuscript preparation: D. Adamopoulos, G. Rovas, N. Johner, N. Stergiopulos, D. Shah. All authors reviewed the results and approved the final version of the manuscript.

* D. Adamopoulos & G. Rovas contributed equally to this study.

## Supplementary Information

Supplementary Information is available for this paper.

