## Supplementary Figures 1-3 for "Left atrial wall shear stress distribution correlates with atrial endocardial electrogram voltage and fibrosis in patients with atrial fibrillation"

**Supplementary Results**

**Supplementary Figure 1.** Pearson's correlation coefficient heatmap between the bipolar voltage and the calculated hemodynamic indices of each subject. The 3D electro-anatomical map of subjects 2, 3, 5, 9, 10, 11 and 14 were obtained in atrial fibrillation.

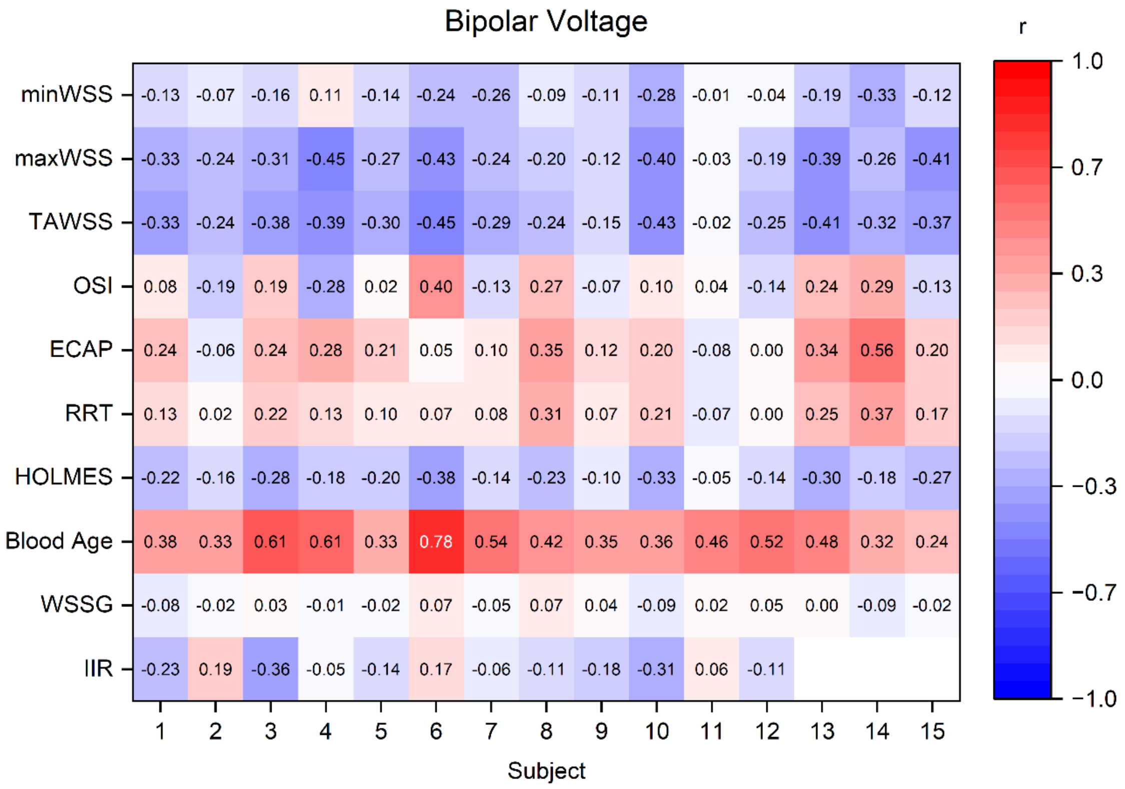

**Supplementary Figure 2.** Pearson's correlation coefficient heatmap between the image intensity ratio and the calculated hemodynamic indices of each subject.

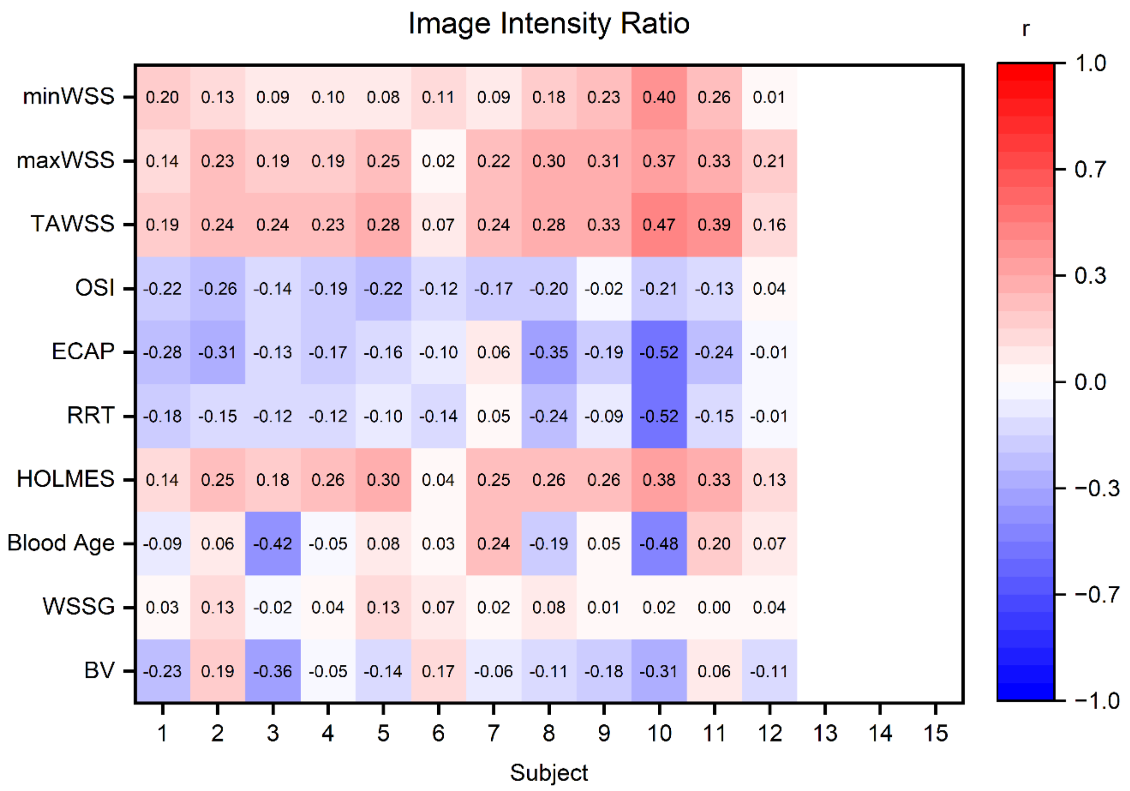

**Supplementary Figure 3.** The 2D maps of the distribution of the time-averaged wall shear stress (TAWSS), bipolar voltage (BV), blood age (BA) and Image Intensity Ratio (IIR) for each subject. The colormap corresponds to healthy regions (black-gray), regions with interstitial fibrosis (white) and fibrotic regions (red). LGE-CMR acquisitions were not available for subjects 13–15.

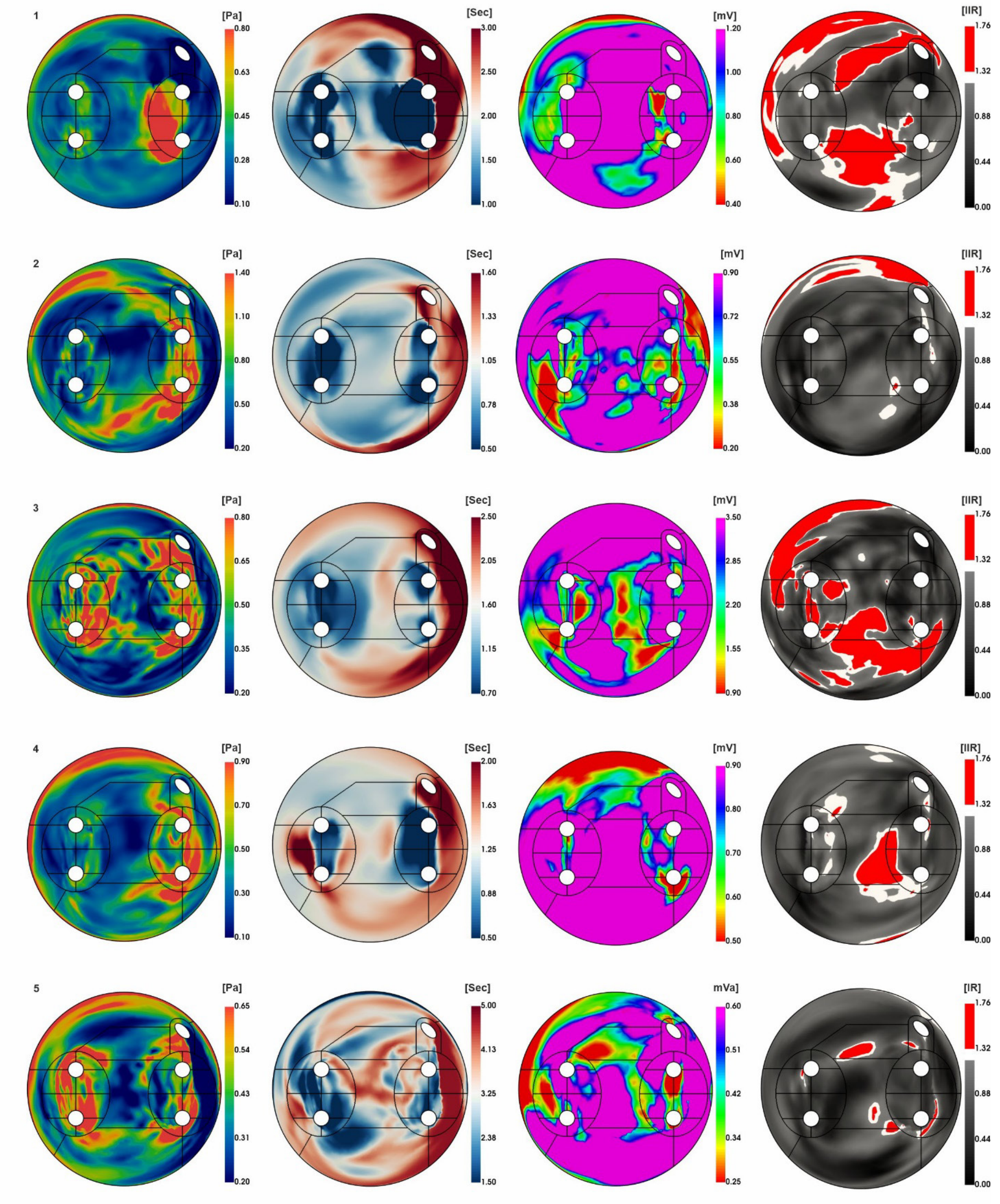

**Supplementary Figure 3** *(continued).* The 2D maps of the distribution of the time-averaged wall shear stress (TAWSS), bipolar voltage (BV), blood age (BA) and Image Intensity Ratio (IIR) for each subject. The colormap corresponds to healthy regions (black-gray), regions with interstitial fibrosis (white) and fibrotic regions (red). LGE-CMR acquisitions were not available for subjects 13–15.

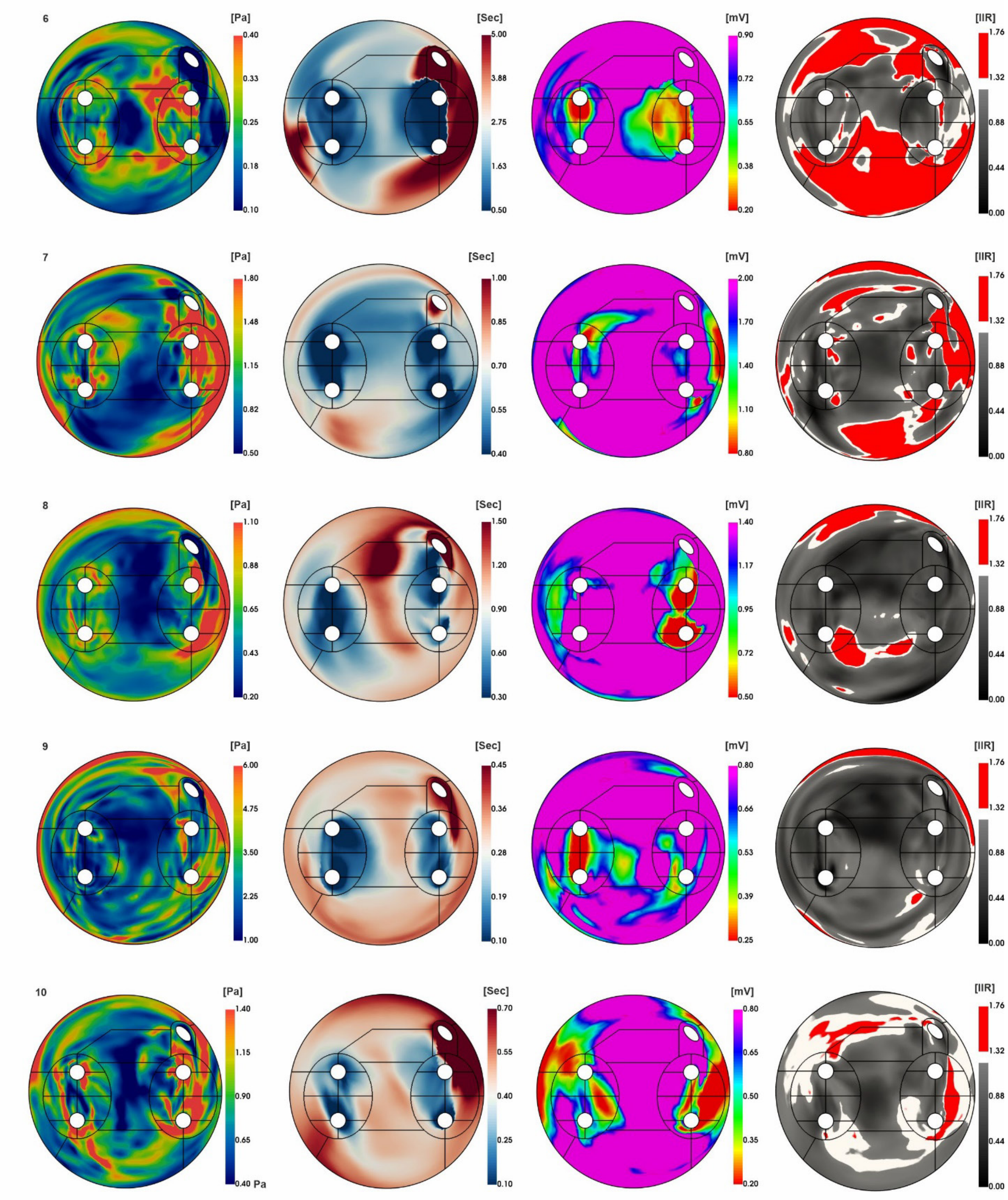

**Supplementary Figure 3** *(continued).* The 2D maps of the distribution of the time-averaged wall shear stress (TAWSS), bipolar voltage (BV), blood age (BA) and Image Intensity Ratio (IIR) for each subject. The colormap corresponds to healthy regions (black-gray), regions with interstitial fibrosis (white) and fibrotic regions (red). LGE-CMR acquisitions were not available for subjects 13–15.

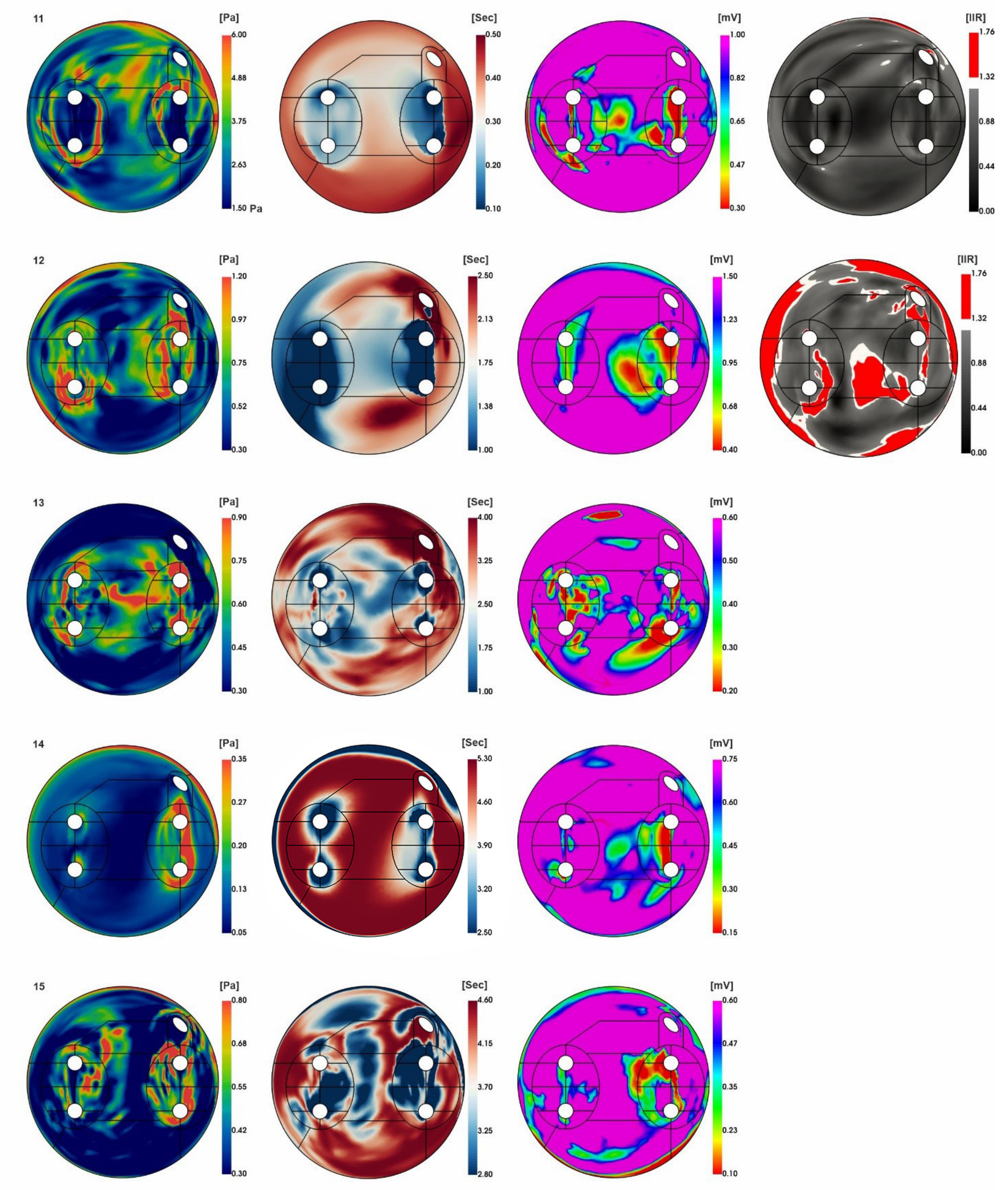
